# Population mortality impacts of the rising cost of living in Scotland: modelling study

**DOI:** 10.1101/2022.11.30.22282579

**Authors:** Elizabeth Richardson, Gerry McCartney, Martin Taulbut, Margaret Douglas, Neil Craig

**Affiliations:** Public Health Scotland, Gyle Square, 1 South Gyle Crescent, Edinburgh, Scotland, EH12 9EB, UK; College of Social Sciences, Adam Smith Building, 28 Bute Gardens, University of Glasgow, Glasgow, G12 8RS, UK; Public Health Scotland, Meridian Court, 5 Cadogan St, Glasgow, G2 6QQ, UK; School of Health & Wellbeing, Glasgow University; Evidence for Action and Public Health Observatory teams, Public Health Scotland, Gyle Square, 1 South Gyle Crescent, Edinburgh, Scotland, EH12 9EB, UK

**Author notes:** Correspondence to: Margaret Douglas, Public Health Scotland, Gyle Square, 1 South Gyle Crescent, Edinburgh, Scotland, EH12 9EB.

## Abstract

**Objectives:** To estimate the potential impacts of unmitigated and mitigated cost of living increases on real household income, mortality, and mortality inequalities in Scotland.

**Design:** Modelling study.

**Setting:** Scotland, 2022/23.

**Participants:** A representative sample of 5,602 Scottish individuals (within 2,704 households) in the 2015/16 Family Resources Survey. We estimated changes in real household income associated with differential price inflation (based on proportion of household spending on different goods and services, by income group), both with and without mitigating UK Government policies, and scaled these to the Scottish population. We estimated mortality effects using a cross-sectional relationship between household income and mortality data, by deprivation group.

**Interventions:** Baseline was Scotland in 2022/23 with the average wage and price inflation of preceding years. The comparison scenarios were unmitigated cost of living increases, and mitigation by the UK Government’s Energy Price Guarantee (EPG) and Cost of Living Support payments.

**Main outcome measures:** Premature mortality rate and life expectancy at birth by Scottish Index of Multiple Deprivation (SIMD) group, and inequalities in both.

**Results:** Unmitigated price inflation was 14.9% for the highest income group and 22.9% for the lowest. UK Government policies partially mitigated impacts of the rising cost of living on real incomes, although households in the most deprived areas of Scotland would still be £1,400 per year worse off than at baseline. With the mitigating measures in place, premature mortality was estimated to increase by up to 6.4%, and life expectancy to decrease by up to 0.9%. Effects would be greater in more deprived areas, and inequalities would increase as a result.

**Conclusions:** Large and inequitable impacts on mortality in Scotland are predicted if real-terms income reductions are sustained. Progressive Cost of Living Support payments are not sufficient to offset the mortality impacts of the greater real income reductions in deprived areas.

**What is already known on this topic:**

- Over the last decade, life expectancy in Scotland has stalled and inequalities have increased.
- Income reductions have been related to increased mortality risk, hence the economy matters for public health.
- The impacts of the rising cost of living and mitigating policies on mortality and inequalities require estimation to inform policymaking.

**What this study adds:**

- The mortality impacts of inflation and real-terms income reduction are likely to be large and negative, with marked inequalities in how these are experienced.
- Current public policy responses are not sufficient to protect health and prevent widening inequalities.
- Bolder and more progressive policy responses are required if health is to improve and health inequalities are to narrow.

## Introduction

After a prolonged period of low inflation, prices have risen rapidly in the UK (and other countries) during 2021 and 2022.^1^ The increase is likely to be due to a combination of factors.^2^ First, the conflict between Russia and Ukraine, and associated sanctions, has restricted the supply of oil and gas, and disrupted the supply of a wide range of food items (including wheat and cooking oil).^2^ The dependence of modern economies on fossil fuels, including for the production and distribution of food, and the consequences on health of restricted availability, has been discussed in detail elsewhere.^3,4^ Second, as the economy has opened after the COVID-19 restrictions, suppliers have had difficulty in meeting the large and rapid increases in demand.^2^ This is in part due to Brexit, which has reduced the supply of labour (with consequent impacts on the ability of producers and suppliers to meet demand).^5^ There is little evidence that inflation has been due to wage bargaining even with this restricted supply of labour, given that wage growth has been substantially below price increases.^1,2^ It is unclear how high inflation will rise, or for how long, not least because of the ongoing UK and geopolitical uncertainties.^1^

The last time the UK experienced inflation of this magnitude was during the 1970s. Although that inflation also had its roots in a fossil fuel supply shock, the impacts are likely to be very different given the profoundly different nature of the economy today.^6^ The current period of inflation and potential impacts on mortality also needs to be put into the context of the longer-term mortality trends. Inequalities in mortality in Scotland have been increasing on most measures since at least the early 1980s.^7^ Average mortality rates, however, had been improving until around 2012, after which the improvement stalled due largely to the impacts of a decade of austerity policies.^8,9^ The COVID-19 pandemic then caused a large increase in mortality in 2020-21 as well as a substantial backlog of unmet healthcare needs.^9,10^ Taken together, this period of inflation is occurring at a particularly perilous time.

The impacts on the affordability of goods and services for large parts of the population, and the consequent impacts on health and health inequalities, are potentially large and worrying.^11,12,13^ Energy costs account for a larger share of the expenditures of poorer households than richer households,^14^ hence energy price hikes will disproportionately affect those on lower incomes.^15^ In mitigation, the UK Government has introduced a universal Energy Price Guarantee (EPG)^16^ and a range of partially targeted Cost of Living Support payments (Box 1).^17,18^ Estimating the population health impacts of unmitigated and mitigated inflation can help inform policymaking. We modelled the possible scale and direction of mortality changes as a result of inflation in Scotland.

### Box 1

**UK Government Cost of Living Support package**

In February and May 2022 the UK Government announced a range of payments to help households with the cost of living crisis:^17,18^

- £400 grant for all domestic energy customers.
- £150 council tax rebate for households in bands A to D (the lowest four of eight house valuation bands), or in receipt of Council Tax Reduction benefit.
- £650 Cost of Living Payment to households in receipt of means-tested benefits.
- £300 Pensioner Cost of Living Payment to pensioner households in receipt of Winter Fuel Allowance/Payment (payable to all of pensionable age).
- £150 Disability Cost of Living Payment to recipients of disability-related benefits.

Household Support Fund monies were also provided to the Scottish Government to enable them to provide directed support as required. This amounted to £41 million for March to September 2022.^18^

## Methods

We modelled how unmitigated and mitigated price inflation would affect real household incomes in 2022/23, and then modelled how this would affect premature mortality, life expectancy, and inequalities in these in Scotland, relative to a ‘business as usual’ baseline. Full details of the modelling are given in the supplementary material, and are summarised here.

### Change in nominal household incomes

First, we used a tax-benefit microsimulation model (UKMOD)^19^ to estimate nominal equivalised household income for Scottish households in the 2015/16 Family Resources Survey (FRS),^20^ for three alternative versions of 2022/23:

1. Average wage inflation for the preceding 10 years (2.6%).^21^
2. Most recent wage inflation forecast for 2022/23 (5.1%).^22^
3. Most recent wage inflation forecast for 2022/23 plus the UK Government’s Cost of Living Support package (Box 1, with the exception of the Household Support Fund, which could not be included because its direct impact on household incomes was not quantifiable).

### Differential change in price inflation

Second, we estimated how price inflation would vary between income quintiles (fifths). Electricity and gas prices have increased at a higher rate than other goods and services,^23^ and the poorest households spend a much bigger share of their total expenditure on electricity and gas than the richest households (supplementary material Table S1)^14^: as a result the price inflation rate experienced by poorer households will be higher than for richer households.

We estimated differential price inflation on the basis of the proportion of household expenditure spent on energy and other goods and services, by income quintile. Annual unmitigated inflation for electricity and gas in October 2022 was calculated between the Office of Gas and Electricity Markets (Ofgem) price caps for October 2021 (costing £1,277 per year for a typical household)^24^ and October 2022 (£3,549 per year for a typical household)^25^: 178%. The UK Government implemented the Energy Price Guarantee of £2,500 per year for a typical household from October 2022,^16^ reducing the annual electricity and gas inflation rate to 96% (i.e., mitigated price inflation for electricity and gas).

We inflated each income quintile’s average expenditure on electricity and gas (Table S1) by our unmitigated and mitigated inflation rate estimates. We inflated their expenditures on other groups by the latest available (August 2022) detailed Consumer Price Index (CPI) annual inflation rates.^23^ Annual inflation in overall household expenditure was then calculated for each income quintile.

### Real income change

Third, we estimated the combined effect of wage inflation, the Cost of Living Support payments, and price inflation on the spending power (real income) of the households in each income quintile in 2022/23. Four scenarios were modelled:

1. ‘Baseline’: 2022/23 if households had experienced the average wage and price inflation rates of the preceding 10 years (2.6%^21^ and 1.8%^26^ respectively).
2. ‘Unmitigated inflation’: 2022/23 with 5.1% wage inflation^22^ and differential unmitigated price inflation (incorporating the £3,549 energy price cap).^25^
3. ‘Mitigated by EPG’: 2022/23 with 5.1% wage inflation and differential mitigated price inflation (incorporating the £2,500 EPG).^16^
4. ‘Mitigated by EPG and Cost of Living Support’: 2022/23 with 5.1% wage inflation, differential price inflation mitigated by the £2,500 EPG, and Cost of Living (COL) Support payments (Box 1).^16,17,18^

We grouped households into income quintiles on the basis of their equivalised income at baseline, using FRS survey weights to produce a distribution that was representative of the Scottish population. Real income was estimated for each household under each scenario, by applying the relevant price inflation rate to the relevant nominal income:

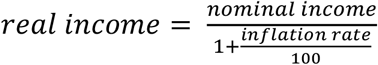

Average real household income under each scenario was then calculated for area deprivation quintiles, using a mapping of the FRS households to Scottish Index of Multiple Deprivation (SIMD) 2016 quintiles, and FRS survey weights. The Gini coefficient was used to estimate inequality in the distribution of real income across the population under each scenario.^27^

### Mortality effect

We used the existing Scottish policy scenario modelling approach – Informing Interventions to reduce health Inequalities (‘Triple I’) – to estimate mortality impacts under each scenario.^28,29,30^ There is evidence that income decline is related to increased mortality risk, but no generalisable effect size is available.^31^ In the absence of synthesised empirical evidence of the relationship between income change and mortality Triple I uses the relationship between income difference and mortality, from cross-sectional data (see supplementary material). As the mortality effect size calculation is a strong assumption, we conducted a sensitivity analysis by reducing the effect by 50%.

The deaths data for each scenario, by age group, sex and SIMD quintile, were then used to calculate summary measures of mortality and inequality: life expectancy (Chiang method),^32^ premature mortality rate (deaths under 75 years), the Slope Index of Inequality (SII, linear) and Relative Index of Inequality (RII, linear).^33^

### Fiscal costs

Direct fiscal costs on payments to households (Cost of Living Support payments and social security) and revenues derived from households (taxes and National Insurance Contributions (NIC)) were estimated for each scenario using UKMOD, and grossed up to the Scottish population using FRS survey weights. The cost of the EPG cannot be estimated from UKMOD, but a crude estimate was obtained by applying Scotland’s population share of Great Britain (8.39% in 2021)^34^ to the UK Government’s estimated cost of the EPG (£31 billion in 2022/23)^35^.

## Results

We estimated that unmitigated inflation in October 2022 would have ranged from 14.9% in the highest income quintile to 22.9% in the lowest income quintile; the EPG reduced this to between 11.7% and 15.7%, respectively (Figure 1).

**Figure 1.**
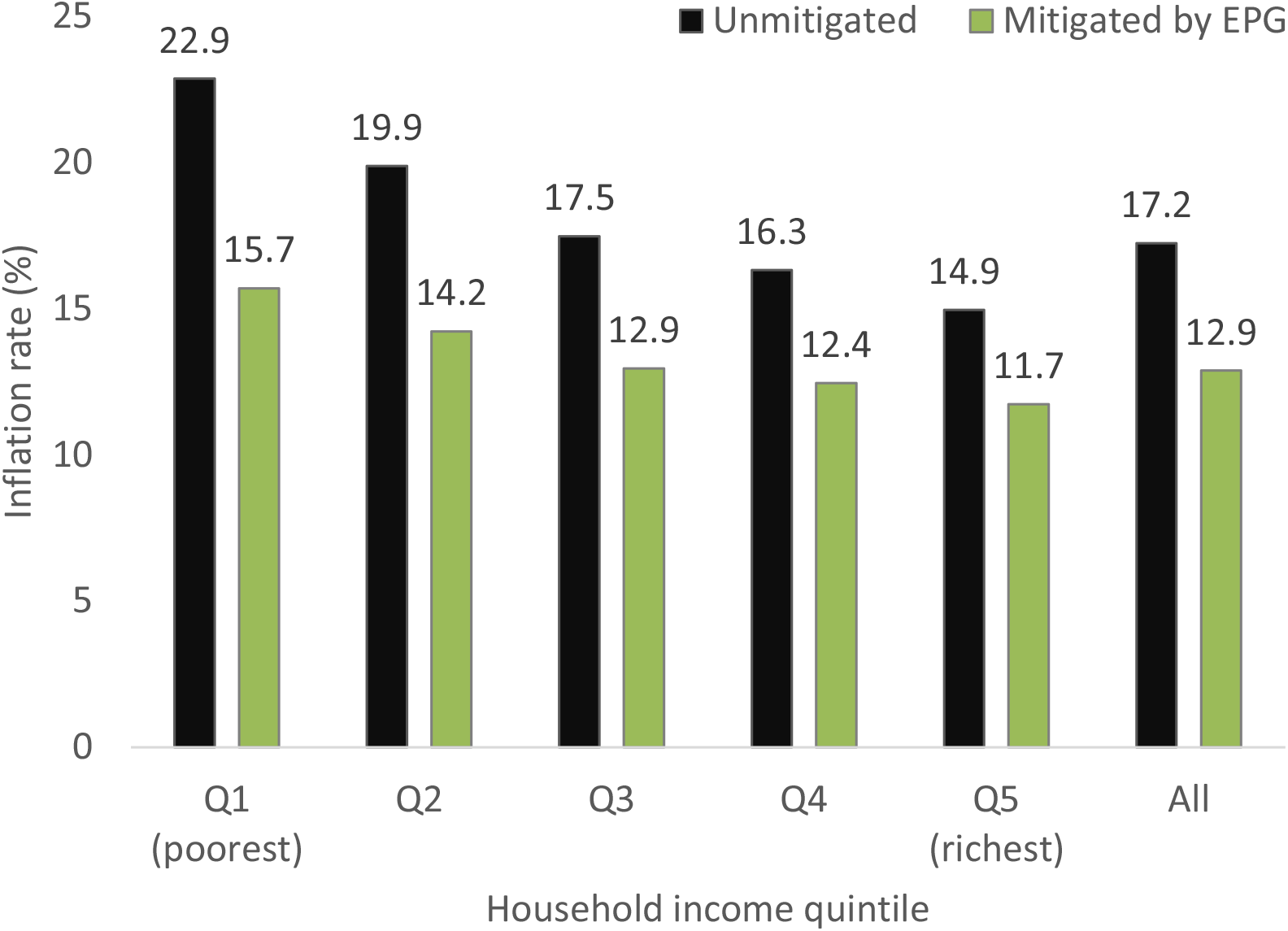
Estimated inflation by UK household income quintile, October 2022.

Real-terms decreases in income would be larger in absolute terms for households in less deprived than more deprived areas under all scenarios (Tables 1 and S4). However, households in the most deprived areas would be hit hardest in relative terms under the ‘Unmitigated inflation’ and ‘Mitigated by EPG’ scenarios. The addition of the partially targeted Cost of Living Support would reverse this, resulting in a smaller relative real-terms drop for those in more deprived than less deprived areas.

**Table 1.** Estimated impact of cost of living scenarios on real incomes in 2022/23, by SIMD 2016 quintile.

| Scenario | SIMD 2016 quintile |  |  |  |  |
| --- | --- | --- | --- | --- | --- |
|  | Q1 (most deprived) | Q2 | Q3 | Q4 | Q5 (least deprived) |
| Baseline mean real household income (£/year) | 25,945 | 29,155 | 31,288 | 34,510 | 42,018 |
| Absolute change (£/year): |  |  |  |  |  |
| Unmitigated | -3,316 | -3,542 | -3,708 | -3,998 | -4,645 |
| Mitigated by EPG | -2,378 | -2,551 | -2,675 | -2,899 | -3,389 |
| Mitigated by EPG and Cost of Living Support | -1,366 | -1,607 | -1,865 | -2,152 | -2,750 |
| Relative change (%): |  |  |  |  |  |
| Unmitigated | -12.8 | -12.1 | -11.9 | -11.6 | -11.1 |
| Mitigated by EPG | -9.2 | -8.7 | -8.5 | -8.4 | -8.1 |
| Mitigated by EPG and Cost of Living Support | -5.3 | -5.5 | -6.0 | -6.2 | -6.5 |

The Gini coefficient for the distribution of real income across the population would be 30.5% at baseline (0 indicates perfect equality while 100 indicates maximum inequality). Income inequality would increase by 4% under ‘Unmitigated inflation’ (Gini 31.8%) and 3% under ‘Mitigated by EPG’ (Gini 31.3%). The addition of Cost of Living Support would reduce income inequality by 2% (Gini 29.9%).

Direct fiscal costs and revenues arising from the scenarios, relative to baseline, are shown in Figure 2. For Scottish households, we estimated that the EPG would cost around £2.6 billion, and the Cost of Living Support payments around £2.2 billion. Compared to baseline, revenue from income tax and National Insurance Contributions (NIC) would increase by around £1.0 billion (3.0%) in all scenarios due to the higher rate of wage inflation, and £0.06 billion less would be spent on pre-existing benefits, due to increased wages. As a result, net fiscal impacts relative to the baseline scenario would be an additional spend of £1.6 billion under ‘Mitigated by EPG’ and £3.7 billion under ‘Mitigated by EPG and Cost of Living Support’.

**Figure 2.**
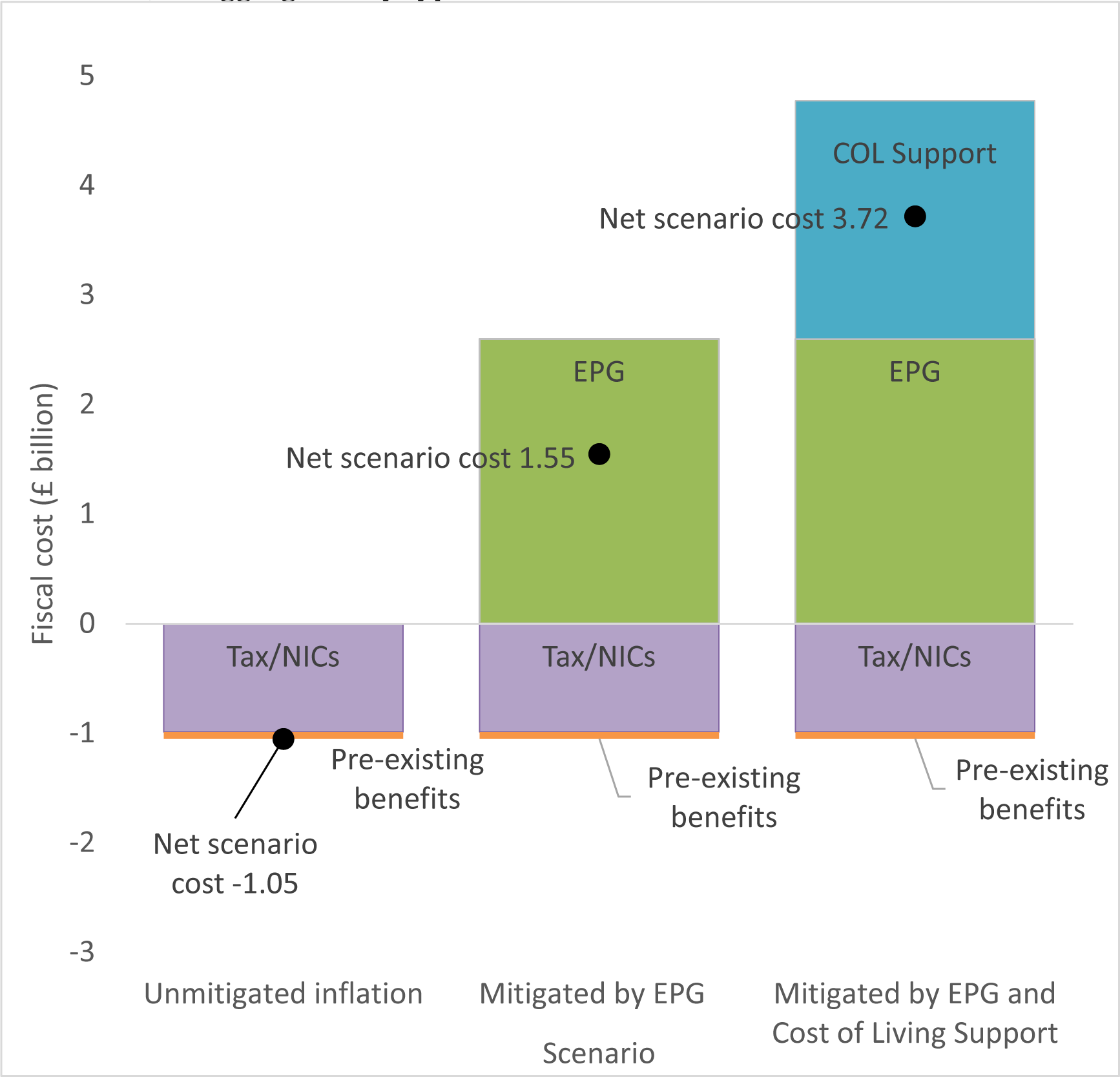
Estimated net fiscal cost of each scenario (£ billion, 2022/23), relative to baseline, disaggregated by type of cost or revenue.

Our model estimated large effects on mortality rates under each scenario, due to their real-terms income decreases: ‘Unmitigated inflation’ was predicted to increase rates by 5% in the least deprived areas and 23% in the most deprived areas (Table S5). Mortality effects would reduce to between 3% and 16% under ‘Mitigated by EPG’ and by between 2% and 8% under ‘Mitigated by EPG and Cost of Living Support’.

Across the population, ‘Unmitigated inflation’ was estimated to increase premature mortality by 16% compared with baseline (Figure 3, Table S6), although effects would be greatest in the most deprived areas (192 more premature deaths per 100,000 population per year: a 23% increase) and smallest in the least deprived (11 more per 100,000 population per year: a 4% increase). The EPG would mitigate the increase somewhat, although premature mortality would still be 15% higher than baseline in the most deprived areas and 3% higher in the least deprived. The stark deprivation gradient in premature mortality change is lessened slightly by the addition of Cost of Living Support, but the most deprived areas are still predicted to experience a relative increase four times larger than the least deprived areas under this scenario. In absolute terms, the income reduction estimated for this scenario could result in 68 more premature deaths per 100,000 population per year in the most deprived areas (6 more per 100,000 population per year in the least deprived).

**Figure 3.**
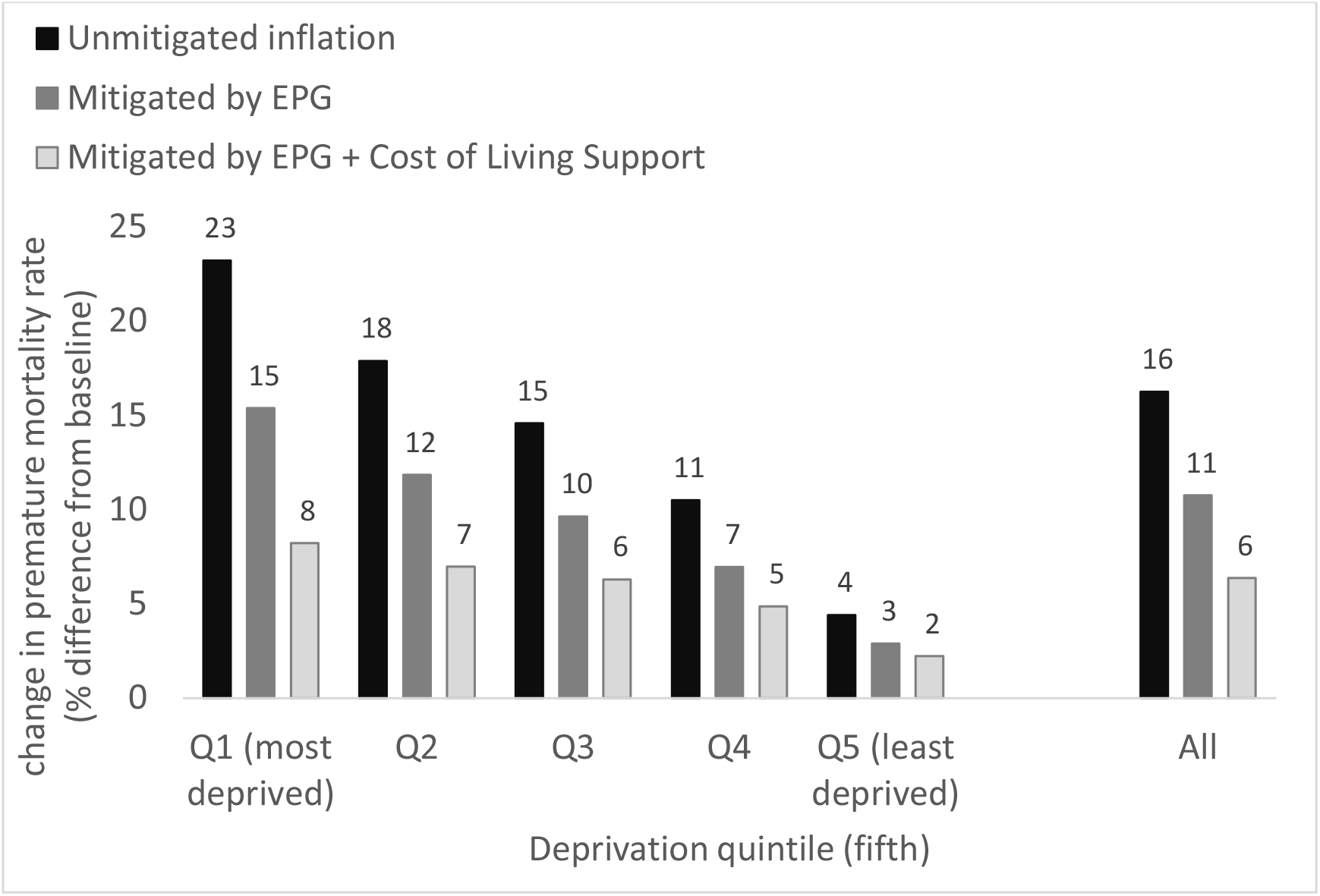
Relative impact of cost of living scenarios on premature mortality rates, compared with the baseline scenario, by SIMD 2016 quintile.

Increases in mortality result in decreased life expectancy. ‘Unmitigated inflation’ was estimated to decrease population-level life expectancy by 2.1% (1.6 years; Figure 4, Table S7). ‘Mitigated by EPG’ would result in a 1.4% fall in life expectancy (1.1 years), and the additional mitigation by Cost of Living Support would reduce the fall to 0.9% (0.7 years). Under each scenario the biggest life expectancy reductions would be experienced in the most deprived areas of Scotland, ranging from 2.7 years (3.7%) under the unmitigated scenario to 1.0 years (1.4%) once mitigation by the EPG and Cost of Living Support payments were included.

**Figure 4.**
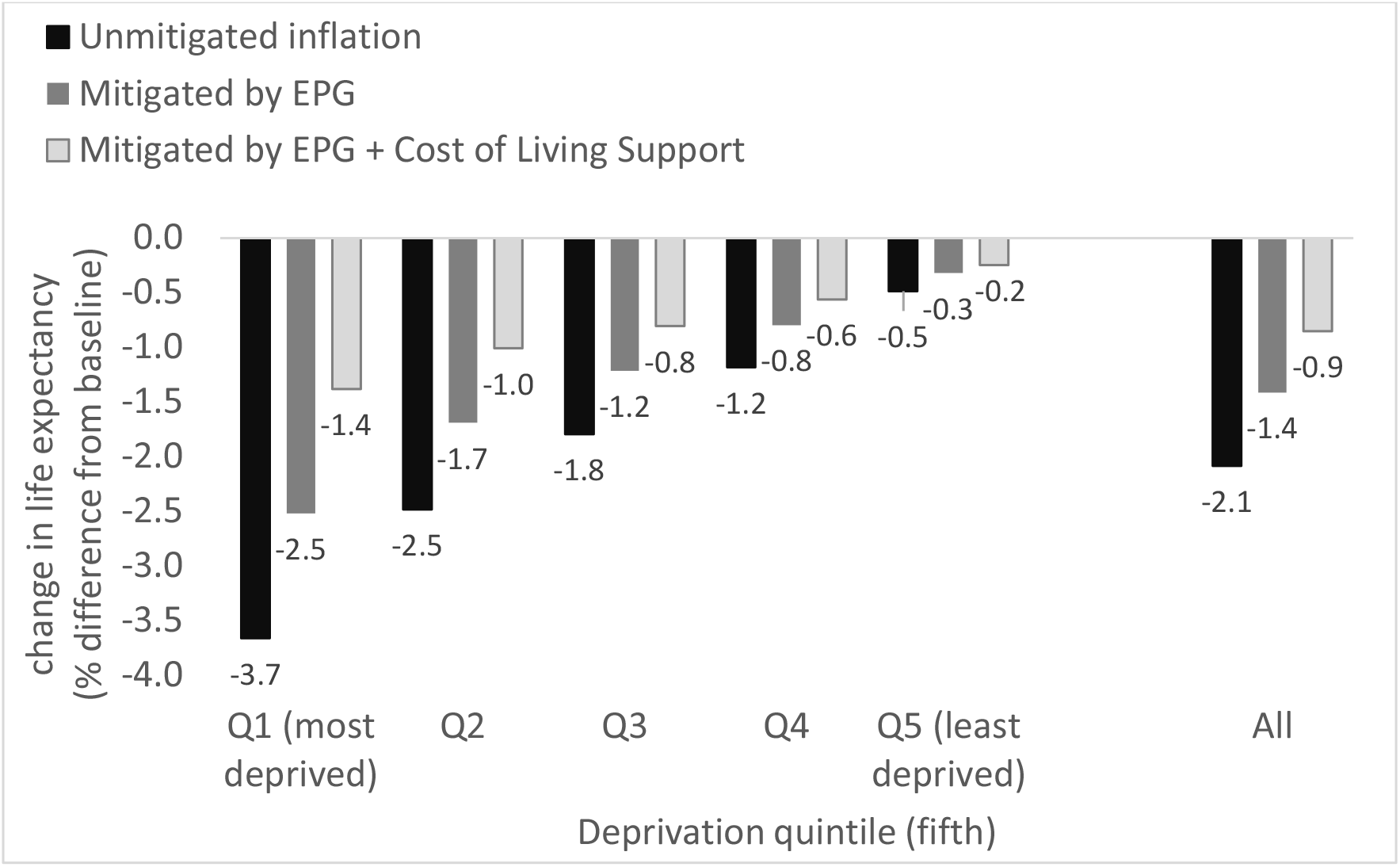
Relative impact of cost of living scenarios on life expectancy at birth, compared with the baseline scenario, by SIMD 2016 quintile.

An alternative way to conceptualise how reductions in real incomes would affect households at different levels of deprivation is presented in Tables 2 and S8. We estimated the change in premature mortality (Table 2) and life expectancy (Table S8) that could result from a range of real income reductions (from 2% to 12%). The results show that the same percentage reduction in real household income would have a much bigger detrimental impact on mortality outcomes in more deprived areas than in less deprived areas. The relative increase in premature mortality in the most deprived quintile would be around five times that in the least deprived quintile, for the same real income reduction (seven times as high for life expectancy). Put another way, to match the premature mortality impact of a 2% real-terms reduction in income in the most deprived areas, incomes in the least deprived areas would need a real-terms reduction of between 8% and 10%.

**Table 2.**
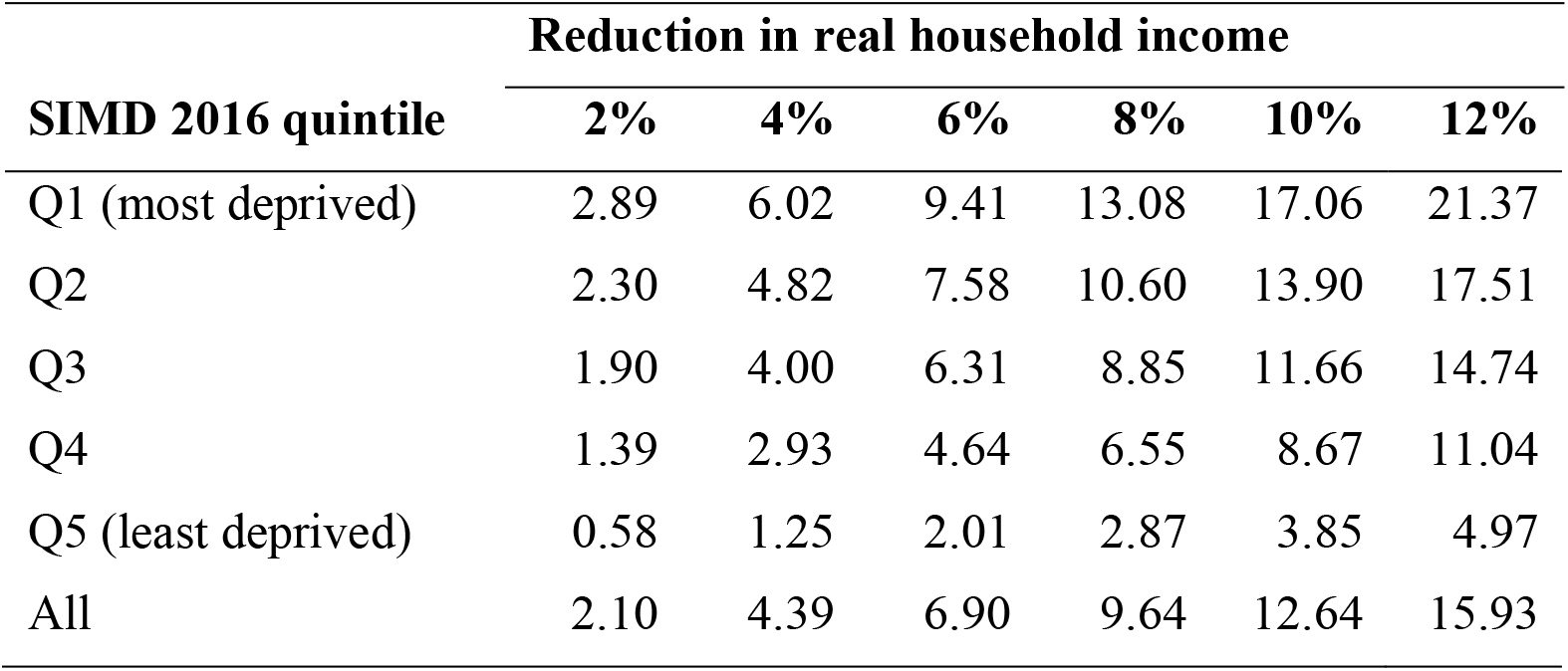
Estimated increase (%) in premature mortality rates compared with baseline for a range of reductions in real household income, by SIMD 2016 quintile.

| SIMD 2016 quintile | Reduction in real household income |  |  |  |  |  |
| --- | --- | --- | --- | --- | --- | --- |
|  | 2% | 4% | 6% | 8% | 10% | 12% |
| Q1 (most deprived) | 2.89 | 6.02 | 9.41 | 13.08 | 17.06 | 21.37 |
| Q2 | 2.30 | 4.82 | 7.58 | 10.60 | 13.90 | 17.51 |
| Q3 | 1.90 | 4.00 | 6.31 | 8.85 | 11.66 | 14.74 |
| Q4 | 1.39 | 2.93 | 4.64 | 6.55 | 8.67 | 11.04 |
| Q5 (least deprived) | 0.58 | 1.25 | 2.01 | 2.87 | 3.85 | 4.97 |
| All | 2.10 | 4.39 | 6.90 | 9.64 | 12.64 | 15.93 |

The Slope Index of Inequality (SII) and Relative Index of Inequality (RII) estimate inequality across the full population, not just between the most and least deprived groups. At baseline, the absolute difference between the most and the least deprived ends of the socioeconomic gradient (SII) was estimated to be 713 additional premature deaths per 100,000 population per year, and 13 fewer years of life expectancy (Table S9). In relative terms, the most deprived end of the socioeconomic gradient had a premature mortality rate 77% higher than the population average, and a life expectancy 8% lower than the population average.

The ‘Unmitigated inflation’ scenario would have increased these pre-existing inequalities considerably: absolute inequalities would increase by 30% for premature mortality and 21% for life expectancy, and relative inequalities would increase by 12% and 23%, respectively. Mitigation by the EPG and Cost of Living Support would lessen these increases, but both absolute and relative inequalities would rise under any scenario modelled here.

To check the sensitivity of the results to the strength of the income-mortality relationship we reduced the size of the effect by 50% for each scenario. The size of each scenario’s effects on life expectancy, premature mortality, and inequalities in both, were reduced by around 50%.

## Discussion

### Principal findings

The population is experiencing high price inflation rates; poorer households are experiencing the highest rates, as they spend a greater proportion of their income on energy. Households in the most deprived areas of Scotland would have seen the biggest relative reduction in income in real terms, but the partially targeted Cost of Living Support payments will give some protection. The payments would partially mitigate the impacts of price increases for households in the most deprived areas, although we estimated they would still be around £1,400 (5.3%) worse off in real terms in 2022/23. Without the Cost of Living Support payments we estimated that income inequality would widen; the addition of Cost of Living Support would reduce income inequality slightly.

Our modelling estimated large and inequitable increases in mortality. Even with mitigation by the EPG and Cost of Living Support payments, real-terms income reductions could result in population-wide premature mortality increases of up to 6.4%, and life expectancy decreases of up to 0.9%. To put this in context, premature mortality in Scotland increased by 7.4% between 2019 and 2020 – an effect that has been largely attributed to COVID-19 deaths.^36^ The effects would be greatest in the most deprived areas, so absolute and relative mortality inequalities would increase.

The Cost of Living Support payments are progressive in that they help to reduce the real income hit experienced by households in the most deprived areas. However, we found that these payments will not be sufficient to offset the potential impacts on mortality of the greater falls in real incomes for these households, and to prevent inequalities from widening, in part because the health impacts of a given change in income are greater for lower income groups.

### Strengths and weaknesses

We employed two well-documented and widely-used policy modelling approaches: UKMOD and Triple I.^19,28^ However, there are several limitations to the methods employed here.

Our price inflation estimates did not include owner-occupiers’ housing costs, or other factors affecting a household’s expenditures. For example, households in Scotland spend more on electricity and gas than the UK average,^37^ so our UK-based rates may be underestimates. Our rates exceed the CPI because ONS downweights electricity and gas expenditure (3.4%), whereas it averaged 5.3% for households in 2021.^14^ Our rates exceed Institute for Fiscal Studies’ estimates due to differing data and time periods.^15^

Our UKMOD modelling assumed uniform wage inflation, although wage growth will differ between public and private sectors. Under-representation of certain groups in the FRS data may affect the accuracy of our calculations. We included policies with a direct quantifiable impact on household incomes, but could not include policies such as the UK Government’s Household Support Fund or the Scottish Government’s rent freeze and eviction ban. Income effects for income quintiles were mapped to deprivation quintiles to enable linkage to routine mortality data; this will have reduced the effect range as there are low and high income households in each deprivation quintile (Table S2).

Estimating the mortality impact of reduced real incomes is difficult and requires numerous assumptions. In the absence of longitudinal evidence, our Triple I modelling used the relationship between income difference and mortality from cross-sectional data. This captures the effects of long-term exposure to different income levels, rather than short-term effects. As such, the mortality impact estimates presented here should not be seen as predictions for 2022/23 in isolation, but rather as an indication of the potential magnitude of effects if income changes are sustained. It is worth noting, however, that over the period 2010/11-2021/22, the Triple I model predicted impacts of changes in income on mortality that were consistent with the pre-pandemic decline in life expectancy in Great Britain.^30,38^ The level of change reported here is therefore plausible if income effects persist over a number of years.

The mortality effects presented do not account for potential behavioural responses to real-terms income reductions – such as working longer hours or reducing energy consumption – which may mitigate economic exposures. Effects of recession, austerity or unemployment were also not considered and may compound the effects. Mortality effects are important to measure, but they are a narrow measure of health; a concurrent health impact assessment considers wider health effects.

### Implications

The economy, and broader political economy, matters for population health.^39^ Evidence suggests that economic conditions in the UK have stalled life expectancy trends and widened health inequalities since around 2012. Austerity has caused real-terms income reductions for the poorest households, and a fraying of the social security system.^9^

The potential impacts of inflation and reduced real incomes are likely to be large and negative, with marked inequalities in how these are experienced. More progressive policy responses are required to protect health and prevent widening inequalities as a result of the Cost of Living Crisis.

## Data Availability

All data produced in the present study are available upon reasonable request to the authors.

## Acknowledgements

We are grateful to Peter Levell, Institute for Fiscal Studies, for advice on the inflation modelling, and Rachel Thomson, University of Glasgow, for advice on the tax-benefit microsimulation. The results presented here are based on UKMOD version A3.23+. UKMOD is maintained, developed and managed by the Centre for Microsimulation and Policy Analysis (CeMPA) at the University of Essex. The process of extending and updating UKMOD is financially supported by the Nuffield Foundation (2018-2021). The results and their interpretation are the authors’ sole responsibility.

## Ethical approval

The study was conducted on anonymised survey data obtained from the UK Data Service under an End User Licence Agreement with the University of Essex. Ethical approval was not required as only secondary and non-identifiable data were used.

## Data availability statement

No additional data available.

## Contributors

GM and ER designed the study. ER conducted the analyses. All authors had access to the study data, contributed to drafting the paper, provided substantive comments on the paper and approved the final version. The corresponding author attests that all listed authors meet authorship criteria and that no others meeting the criteria have been omitted. All authors approved the decision to submit for publication. ER is the guarantor.

## Transparency statement

The lead author (the manuscript’s guarantor) affirms that the manuscript is an honest, accurate, and transparent account of the study being reported; and that no important aspects of the study have been omitted.

