## Supplemental Information for "Population mortality impacts of the rising cost of living in Scotland: modelling study"

**Abstract**

*Objectives*

To estimate the potential impacts of unmitigated and mitigated cost of living increases on real household income, mortality, and mortality inequalities in Scotland.

*Design*

Modelling study.

*Setting*

Scotland, 2022/23.

*Participants*

A representative sample of 5,602 Scottish individuals (within 2,704 households) in the 2015/16 Family Resources Survey. We estimated changes in real household income associated with differential price inflation (based on proportion of household spending on different goods and services, by income group), both with and without mitigating UK Government policies, and scaled these to the Scottish population. We estimated mortality effects using a cross-sectional relationship between household income and mortality data, by deprivation group.

*Interventions*

Baseline was Scotland in 2022/23 with the average wage and price inflation of preceding years. The comparison scenarios were unmitigated cost of living increases, and mitigation by the UK Government’s Energy Price Guarantee (EPG) and Cost of Living Support payments.

*Main outcome measures*

Premature mortality rate and life expectancy at birth by Scottish Index of Multiple Deprivation (SIMD) group, and inequalities in both.

*Results*

Unmitigated price inflation was 14.9% for the highest income group and 22.9% for the lowest. UK Government policies partially mitigated impacts of the rising cost of living on real incomes, although households in the most deprived areas of Scotland would still be £1,400 per year worse off than at baseline. With the mitigating measures in place, premature mortality was estimated to increase by up to 6.4%, and life expectancy to decrease by up to 0.9%. Effects would be greater in more deprived areas, and inequalities would increase as a result.

*Conclusions*

Large and inequitable impacts on mortality in Scotland are predicted if real-terms income reductions are sustained. Progressive Cost of Living Support payments are not sufficient to offset the mortality impacts of the greater real income reductions in deprived areas.

**Part 1: Detailed methodology**

*Change in household incomes*

To set up our study scenarios we first needed to model changes in nominal income at the household level, before accounting for changes in the spending power of that income. We modelled three versions of 2022/23 in UKMOD version A3.23+^[1]^ – a detailed open-source tax-benefit microsimulation model – using the EUROMOD software (v3.5.1). Version 1 was 2022/23 with the average wage inflation for the preceding 10 years (2.62%).^[2]^ Version 2 included the most recent Office for Budget Responsibility wage inflation forecast for 2022/23 (5.1%).^[3]^ Version 3 included 5.1% wage inflation and the UK Government’s Cost of Living Support package (see Box 1 in main paper). The Household Support Fund was not included because its direct impact on household incomes was not quantifiable.

The effect of these changes to incomes in 2022/23 was modelled for Scottish respondents to the 2015/16 Family Resources Survey (FRS),^[4]^ as the area deprivation level (Scottish Index of Multiple Deprivation 2016) for these respondents was known, and SIMD was required for linking household data to routine health data. Household incomes before housing costs were calculated and equivalised using the Organisation for Economic Co-operation and Development (OECD) modified equivalence scale, in which the reference is a couple with no children. The processing was conducted in RStudio.^[5]^

*Change in price inflation rate*

Price inflation affects how far a household’s nominal income will stretch. Electricity and gas prices have increased at a higher rate than other goods and services,^[6]^ and the poorest households spend a much bigger share of their total expenditure on electricity and gas than the richest households (Table S1 and Figure S1): as a result the price inflation rate experienced by poorer households will be higher than for richer households.

We calculated differential price inflation rates for quintiles (fifths) of the household income distribution as follows. We applied different inflation rates to the household expenditure groups detailed in Table S1. For electricity and gas, Ofgem’s price cap for 1 October 2021 – 31 March 2022 was set at a level that would lead to energy costs of £1,277 per year for an average household.^[7]^ From 1 October 2022 the price cap was set to increase to a level that would lead to energy costs of £3,549 per year for an average household,^[8]^ but the UK Government replaced this with an Energy Price Guarantee to ensure that a typical household spends no more than £2,500 per year.^[9]^ The annual inflation of electricity and gas prices (Oct 2021 to Oct 2022) would therefore have been 177.9% with the original price cap, but was reduced to 95.8% under the EPG. For other fuels (mainly oil and coal), inflation has also been higher than for other expenditure groups: 86.2% for liquid fuels and 29.8% for solid fuels in August 2022.^[10]^ These fuels are aggregated in the household expenditure tables, so we applied their average inflation (unweighted): 58.0%. For the other expenditure groups we applied the latest available consumer price index (CPI) figures (August 2022) for each group.^10^ Inflation for housing expenditures (20.0%) was skewed upwards by energy costs, therefore we used the average for non-energy expenditures (5.0%).

Average household expenditures (Table S1) were inflated by the specified figures, and the resulting overall expenditure was divided by the original expenditure to give an annual inflation figure for each income quintile (Figure S2). We estimated that unmitigated inflation in 2022 would have ranged from 14.9% in the highest income quintile to 22.9% in the lowest income quintile, and that the EPG reduced this to between 11.7% (highest income quintile) and 15.7% (lowest income quintile).

The Institute for Fiscal Studies (IFS) estimated differential price inflation in a similar way in August 2022.^[11]^ The IFS estimated that unmitigated inflation would result in price inflation ranging from 10.9% for the highest income quintile, to 17.6% for the lowest income quintile. Our estimates are higher, which is likely to be explained by the baseline: we used October 2021 while IFS used April 2022. We also used more recent data and more detailed Consumer Price Inflation (CPI) forecasts. We expected our price inflation estimates to be higher than the Office for National Statistics (ONS) official CPI figure because households spend a greater proportion of their total expenditure on these fuels (between 3.9% in richest households and 8.7% in poorest households; Figure S1) than the 3.4% weighting given to them in the CPI calculation.^10^

*Change in real household incomes*

The next step was to model change in spending power, or real income, under each of our study scenarios:

1. ‘Baseline’: 2022 if households had experienced the average wage and price inflation rates of the preceding 10 years (2.62%^[12]^ and 1.81%^[13]^ respectively).
2. ‘Unmitigated inflation’: 2022 with 5.1% wage inflation for all wage-earners and differential price inflation that incorporates the £3,549 price cap (14.9% to 22.9%).
3. ‘Mitigated by EPG’: 2022 with 5.1% wage inflation and differential price inflation mitigated by the £2,500 Energy Price Guarantee (11.7% to 15.7%).
4. ‘Mitigated by EPG and Cost of Living Support’: 2022 with 5.1% wage inflation, differential price inflation mitigated by the £2,500 EPG, and Cost of Living Support payments (Box 1).

We grouped households in the baseline scenario into equivalised household income quintiles, using FRS survey weights to produce a distribution that was representative of the Scottish population. We calculated real income for each household under each scenario using the relevant differential price inflation rate (£3,549 price cap or £2,500 EPG). For example, a household in the poorest income quintile would experience price inflation of 22.9% in the ‘Unmitigated inflation’ scenario, and price inflation of 15.7% in the scenarios involving mitigation by the EPG. Real income was calculated as follows:

$$real income= \frac{nominal income}{1+\frac{inflation rate}{100}}$$

where income is equivalised household nominal income at baseline, and rate is the differential price inflation rate (%). The average annual CPI inflation rate for the preceding 10 years (1.81%)^13^ was applied to all households in the baseline scenario. We used a mapping of Scottish 2015/16 FRS respondents to SIMD 2016 quintiles (Table S2) to calculate average real income in each quintile, under each scenario. The averaging was weighted using survey response weights to ensure representativeness.

*Change in population mortality*

We used the existing Scottish policy scenario modelling approach – Informing Interventions to reduce health Inequalities (‘Triple I’)^[14],^^[15],^^[16]^ – to estimate mortality impacts under each scenario.

National Records of Scotland (NRS) deaths data for 2021, by 5-year age group, SIMD quintile and sex, were used for the baseline scenario.^[17]^ Death counts at baseline were then modified by an estimate of the effect of income change under each scenario on mortality.

In the absence of synthesised empirical evidence of the relationship between income change and mortality we used the relationship between income difference and mortality, from cross-sectional data (Table S3). The choice of function was driven by the asymptotic shape of the plotted data, and the assumption that the same absolute or relative change in income will have a larger effect on health for lower income than higher income households. A non-linear least squares logistic regression model gave a good fit to the data (Figure S2), with the formula:

$$EASR= \frac{1+ e^{\frac{1826.84-income}{539.41}}}{0.00124}$$

Rate ratios for the mortality effect under each scenario, by SIMD quintile, were estimated by dividing the scenario European Age-Standardised Rate (EASR) by the baseline EASR. To estimate deaths occurring under each scenario we multiplied deaths under the baseline scenario by the relevant rate ratio. As the mortality effect size calculation is a strong assumption, we conducted a sensitivity analysis by reducing the effect by 50%.

The deaths data for each scenario, by age group, sex and SIMD quintile, were then used to calculate summary measures of mortality and inequality: life expectancy (Chiang method),^[18]^ premature mortality rate (deaths under 75 years), the Slope Index of Inequality (SII, linear) and Relative Index of Inequality (RII, linear).^[19]^

**Table S1 - Average weekly expenditure (£) on CPI goods and services, by expenditure group and UK income quintile, FY 2020/21.**

| **Group** | **Q1 (poorest)** | **Q2** | **Q3** | **Q4** | **Q5 (richest)** | **All** |
| --- | --- | --- | --- | --- | --- | --- |
| Food and non-alcoholic beverages | 39.65 | 56.90 | 67.20 | 80.25 | 102.10 | 69.20 |
| Alcoholic beverages and tobacco | 8.40 | 10.25 | 13.65 | 16.55 | 21.55 | 14.10 |
| Clothing and footwear | 5.00 | 10.15 | 13.00 | 17.15 | 26.95 | 14.50 |
| Housing, water, and energy (excl. fuel) | 39.25 | 54.15 | 68.70 | 64.55 | 80.40 | 61.40 |
| Electricity and gas | 17.45 | 20.95 | 22.10 | 22.65 | 26.85 | 22.00 |
| Other fuels | 0.65 | 1.05 | 1.15 | 1.45 | 1.60 | 1.20 |
| Household goods and services | 15.15 | 24.40 | 33.85 | 37.90 | 61.05 | 34.47 |
| Health | 2.70 | 5.55 | 6.70 | 9.00 | 9.40 | 6.70 |
| Transport | 17.40 | 35.95 | 54.05 | 80.05 | 116.85 | 60.80 |
| Communication | 12.00 | 16.85 | 21.80 | 24.30 | 29.80 | 20.90 |
| Recreation and culture | 19.25 | 27.95 | 45.15 | 55.20 | 80.10 | 45.50 |
| Education | 0.00 | 2.90 | 3.60 | 5.95 | 27.65 | 8.30 |
| Restaurants and hotels | 5.25 | 9.40 | 15.60 | 23.20 | 38.05 | 18.30 |
| Miscellaneous goods and services | 17.90 | 27.75 | 34.95 | 44.05 | 61.20 | 37.20 |
| **All expenditure groups** | **200.05** | **304.20** | **401.50** | **482.25** | **683.55** | **414.60** |

Source: ONS Family Spending expenditure tables,^[20]^ from Living Costs and Food Survey (Table A6).

**Figure S1. Household expenditure on (a) electricity and gas, and (b) food and non-alcoholic drinks, as a percentage of total expenditure, by UK income quintile, financial year 2020/21.**

% household expenditure

| Household income quintile | Electricity and gas | Food and non-alcoholic drinks |
| --- | --- | --- |
| Q1 (poorest) | 8.72282 | 19.82 |
| Q2 | 6.88692 | 18.7048 |
| Q3 | 5.50436 | 16.7372 |
| Q4 | 4.69673 | 16.6407 |
| Q5 (richest) | 3.92802 | 14.9367 |
| All | 5.30632 | 16.6908 |

Source: ONS Family Spending expenditure tables,^20^ from Living Costs and Food Survey (Table A6).

**Table S2 - Distribution (%) of 2015/16 Family Resources Survey households in each SIMD 2016 quintile between household income quintiles (baseline scenario, 2022 projection).**

|  | **Household income quintile (Scotland-specific)** | | | | |  |
| --- | --- | --- | --- | --- | --- | --- |
| **SIMD 2016 quintile** | **Q1 (poorest)** | **Q2** | **Q3** | **Q4** | **Q5 (richest)** | **Total** |
| Q1 (most deprived) | 29.7 | 25.7 | 19.2 | 15.6 | 9.8 | 100.0 |
| Q2 | 18.2 | 25.5 | 23.3 | 18.4 | 14.6 | 100.0 |
| Q3 | 19.9 | 19.6 | 19.1 | 23.5 | 17.9 | 100.0 |
| Q4 | 18.2 | 14.4 | 21.2 | 22.3 | 23.8 | 100.0 |
| Q5 (least deprived) | 12.0 | 13.5 | 16.4 | 19.8 | 38.4 | 100.0 |

Source: Bespoke linkage of Family Resources Survey 2015/16 Scottish respondents to SIMD 2016 quintiles, conducted by DWP in 2018. Weighted using FRS survey weights. Households assigned to income quintiles (equivalised, before housing costs) under the baseline scenario.

**Table S3 - SIMD quintile-level household income and mortality rate data used to estimate the relationship between income and mortality.**

| **SIMD 2016 quintile** | **Mean equivalised household income before housing costs (£/month), 2015/16** | **Mortality EASR (per 100,000), 2015** |
| --- | --- | --- |
| Q1 (most deprived) | 1,833 | 1,597 |
| Q2 | 2,085 | 1,324 |
| Q3 | 2,269 | 1,172 |
| Q4 | 2,497 | 1,033 |
| Q5 (least deprived) | 3,084 | 890 |

Source: Weighted averages of household income data for 2015/16 Scottish FRS respondents^4^ linked to SIMD 2016 quintiles (DWP mapping); NRS population and deaths data. EASRs standardised to the 2013 European Standard Population.

**Figure S2 – Logistic relationship between household income and mortality rate.**

Red dots show the SIMD-level data points used to fit the model (Table S4), and the black line shows the fitted model predictions.

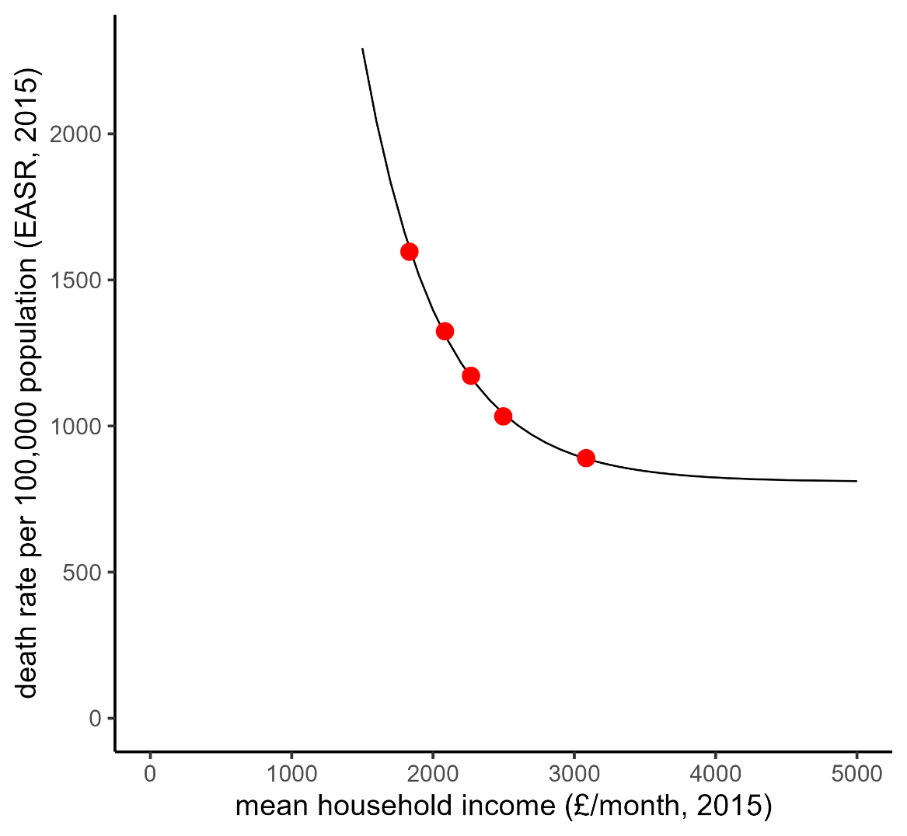

**Part 2: Additional results**

**Table S4 - Estimated mean real household income (£/year, 2022) under each scenario, by SIMD 2016 quintile.**

|  | **SIMD 2016 quintile** | | | | |
| --- | --- | --- | --- | --- | --- |
| **Scenario** | **Q1 (most deprived)** | **Q2** | **Q3** | **Q4** | **Q5 (least deprived)** |
| Baseline | 2,162 | 2,430 | 2,607 | 2,876 | 3,501 |
| Unmitigated inflation | 1,886 | 2,134 | 2,298 | 2,543 | 3,114 |
| Mitigated by EPG | 1,964 | 2,217 | 2,384 | 2,634 | 3,219 |
| Mitigated by EPG and Cost of Living Support | 2,048 | 2,296 | 2,452 | 2,696 | 3,272 |

Projected equivalised real household income before housing costs for 2022 calculated using UKMOD output for FRS 2015/16 respondents. Due to the modelling assumptions and limitations these will differ from actual household income figures for 2022.

**Table S5 - Rate ratios for the mortality rate effect under each scenario compared with baseline, by SIMD 2016 quintile**

|  | **SIMD 2016 quintile** | | | | |
| --- | --- | --- | --- | --- | --- |
| **Scenario** | **Q1 (most deprived)** | **Q2** | **Q3** | **Q4** | **Q5 (least deprived)** |
| Baseline | 1.00 | 1.00 | 1.00 | 1.00 | 1.00 |
| Unmitigated inflation | 1.23 | 1.18 | 1.15 | 1.11 | 1.05 |
| Mitigated by EPG | 1.16 | 1.12 | 1.10 | 1.07 | 1.03 |
| Mitigated by EPG and Cost of Living Support | 1.08 | 1.07 | 1.06 | 1.05 | 1.02 |

**Table S6 – Premature mortality rate per 100,000 per year under each scenario, by SIMD 2016 quintile**

|  | **SIMD 2016 quintile** | | | | |  |
| --- | --- | --- | --- | --- | --- | --- |
| **Scenario** | **Q1 (most deprived)** | **Q2** | **Q3** | **Q4** | **Q5 (least deprived)** | **All** |
| Baseline | 827 | 588 | 408 | 308 | 243 | 463 |
| Unmitigated inflation | 1,019 | 693 | 468 | 341 | 254 | 539 |
| Mitigated by EPG | 955 | 657 | 448 | 330 | 250 | 513 |
| Mitigated by EPG and Cost of Living Support | 895 | 628 | 434 | 323 | 249 | 493 |

European Age-Standardised Rates (EASRs) standardised to the European Standard Population 2013.

**Table S7 – Life expectancy at birth (years) under each scenario, by SIMD 2016 quintile**

|  | **SIMD 2016 quintile** | | | | |  |
| --- | --- | --- | --- | --- | --- | --- |
| **Scenario** | **Q1 (most deprived)** | **Q2** | **Q3** | **Q4** | **Q5 (least deprived)** | **All** |
| Baseline | 72.6 | 76.3 | 79.5 | 81.7 | 83.3 | 78.5 |
| Unmitigated inflation | 69.9 | 74.4 | 78.0 | 80.8 | 82.9 | 76.9 |
| Mitigated by EPG | 70.8 | 75.0 | 78.5 | 81.1 | 83.0 | 77.4 |
| Mitigated by EPG and Cost of Living Support | 71.6 | 75.6 | 78.8 | 81.3 | 83.1 | 77.9 |

**Table S8 - Estimated decrease (%) in life expectancy compared with baseline for a range of reductions in real household income, by SIMD 2016 quintile.**

|  | **Reduction in real household income** | | | | | |
| --- | --- | --- | --- | --- | --- | --- |
| **SIMD 2016 quintile** | **2%** | **4%** | **6%** | **8%** | **10%** | **12%** |
| Q1 (most deprived) | -0.50 | -1.03 | -1.59 | -2.17 | -2.78 | -3.41 |
| Q2 | -0.35 | -0.72 | -1.11 | -1.53 | -1.98 | -2.45 |
| Q3 | -0.25 | -0.52 | -0.81 | -1.13 | -1.47 | -1.83 |
| Q4 | -0.16 | -0.34 | -0.54 | -0.75 | -0.99 | -1.24 |
| Q5 (least deprived) | -0.07 | -0.14 | -0.22 | -0.32 | -0.43 | -0.55 |
| All | -0.28 | -0.59 | -0.92 | -1.27 | -1.64 | -2.04 |

**Table S9 – Effect of each scenario on inequalities in mortality outcomes, compared with baseline.**

|  | **Premature mortality rate** | | **Life expectancy at birth** | |
| --- | --- | --- | --- | --- |
| **Scenario** | **Absolute inequality**  **(linear SII)** | **Relative inequality (linear RII)** | **Absolute inequality**  **(linear SII)** | **Relative inequality (linear RII)** |
| Baseline | 713 per 100,000 population per year | 1.54 | -13.1 years | -0.17 |
| Percent increase from baseline: |  |  |  |  |
| Unmitigated inflation | 30.1% | 11.9% | 20.6% | 23.2% |
| Mitigated by EPG | 20.0% | 8.3% | 14.3% | 15.9% |
| Mitigated by EPG and Cost of Living Support | 10.4% | 3.8% | 7.2% | 8.2% |

Notes: Absolute inequality measured using the Slope Index of Inequality (SII, linear) and relative inequality measured using the Relative Index of Inequality (RII, linear).^19^
